# Autoimmune diseases and risk of non-Hodgkin lymphoma: A Mendelian randomisation study

**DOI:** 10.1101/2024.01.20.24301459

**Authors:** Xiaoting Shi, Joshua D. Wallach, Xiaomei Ma, Tormod Rogne

**Author notes:** **Corresponding author:** Xiaoting Shi, MPhil, Department of Environmental Health Sciences, Yale School of Public Health, New Haven, Connecticut, USA, ORCID number: Xiaoting Shi, Joshua Wallach, Tormod Rogne: Twitter: @Xiaoting_yale, @JoshuaDWallach, @TormodRogne.

## Abstract

**Objective:** To examine whether genetically predicted susceptibility to ten autoimmune diseases (Behçet’s disease, coeliac disease, dermatitis herpetiformis, lupus, psoriasis, rheumatoid arthritis, sarcoidosis, Sjögren’s syndrome, systemic sclerosis, and type 1 diabetes) is associated with risk of non-Hodgkin lymphoma (NHL).

**Design:** Two sample Mendelian randomization (MR) study.

**Setting:** Genome wide association studies (GWASs) of ten autoimmune diseases, NHL, and four NHL subtypes (i.e., follicular lymphoma, mature T/natural killer-cell lymphomas, non-follicular lymphoma, and other and unspecified types of NHL).

**Analysis:** We used data from the largest publicly available GWASs of European ancestry for each autoimmune disease, NHL, and NHL subtypes. For each autoimmune disease, we extracted single nucleotide polymorphisms (SNPs) strongly associated (*P* < 5×10^−8^) with that disease and that were independent of one another (R^2^ < 1×10^−3^) as genetic instruments. SNPs within the human leukocyte antigen region were not considered due to potential pleiotropy. Our primary MR analysis was the inverse-variance weighted analysis. Additionally, we conducted MR-Egger, weighted mode, and weighted median regression to address potential bias due to pleiotropy, and robust adjusted profile scores to address weak instrument bias. We carried out sensitivity analysis limited to the non-immune pathway for nominally significant findings. To account for multiple testing, we set the thresholds for statistical significance at *P* < 5×10^−3^.

**Participants:** The number of cases and controls identified in the relevant GWASs were 437 and 3,325 for Behçet’s disease, 4,918 and 5,684 for coeliac disease, 435 and 341,188 for dermatitis herpetiformis, 4,576 and 8,039 for lupus, 11,988 and 275,335 for psoriasis, 22,350 and 74,823 for rheumatoid arthritis, 3,597 and 337,121 for sarcoidosis, 2,735 and 332,115 for Sjögren’s syndrome, 9,095 and 17,584 for systemic sclerosis, 18,942 and 501,638 for type 1 diabetes, 2,400 and 410,350 for NHL; and 296 to 2,340 cases and 271,463 controls for NHL subtypes. **Exposures**: Genetic variants predicting ten autoimmune diseases: Behçet’s disease, coeliac disease, dermatitis herpetiformis, lupus, psoriasis, rheumatoid arthritis, sarcoidosis, Sjögren’s syndrome, systemic sclerosis, and type 1 diabetes.

**Main outcome measures:** Estimated associations between genetically predicted susceptibility to ten autoimmune diseases and the risk of NHL.

**Results:** The variance of each autoimmune disease explained by the SNPs ranged from 0.3% to 3.1%. Negative associations between type 1 diabetes and sarcoidosis and the risk of NHL were observed (odds ratio [OR] 0.95, 95% confidence interval [CI]: 0.92 to 0.98, *P* = 5×10^-3^, and OR 0.92, 95% CI: 0.85 to 0.99, *P* = 2.8×10^-2^, respectively). These findings were supported by the sensitivity analyses accounting for potential pleiotropy and weak instrument bias. No significant associations were found between the other eight autoimmune diseases and NHL risk. Of the NHL subtypes, type 1 diabetes was most strongly associated with follicular lymphoma (OR 0.91, 95% CI: 0.86 to 0.96, *P* = 1×10^-3^), while sarcoidosis was most strongly associated with other and unspecified NHL (OR 0.86, 95% CI: 0.75 to 0.97, *P* = 1.8×10^-2^).

**Conclusions:** These findings suggest that genetically predicted susceptibility to type 1 diabetes, and to some extent sarcoidosis, might reduce the risk of NHL. However, future studies with different datasets, approaches, and populations are warranted to further examine the potential associations between these autoimmune diseases and the risk of NHL.

**WHAT IS ALREADY KNOWN ON THIS TOPIC:**

1. The etiology of non-Hodgkin lymphoma, a common hematological malignancy, is not fully understood.
2. Observational studies have reported statistically significant associations between ten autoimmune diseases (Behçet’s disease, coeliac disease, dermatitis herpetiformis, lupus, psoriasis, rheumatoid arthritis, sarcoidosis, Sjögren’s syndrome, systemic sclerosis, and type 1 diabetes) and risk of non-Hodgkin lymphoma, but these studies may be susceptible to residual confounding and reverse causation.

**WHAT THIS STUDY ADDS:**

1. Genetically predicted susceptibility to type 1 diabetes, and to some extent sarcoidosis, may be associated with a reduced risk of non-Hodgkin lymphoma, while no clear associations were observed between the other eight autoimmune diseases and risk of non-Hodgkin lymphoma or its subtypes.

**HOW THIS STUDY MIGHT AFFECT RESEARCH, PRACTICE, OR POLICY:**

1. Using an approach that seeks to address residual confounding and reverse causation, these findings contradict previously reported associations between autoimmune diseases and risk of non-Hodgkin lymphoma from traditional observational studies.
2. Future studies with different datasets, approaches, and populations are warranted to further examine the potential associations between these autoimmune diseases and the risk of NHL.

## INTRODUCTION

Non-Hodgkin lymphoma (NHL), a hematological malignancy that arises from lymphocytes, is one of the most common cancers, with more than 544,000 new cases every year worldwide.^1,2^ Despite substantial efforts to identify risk factors for NHL, the exact etiology remains elusive.^3^

According to a recent umbrella review (i.e., systematic review of meta-analyses) evaluating the associations between 134 unique environmental risk factors and the risk of NHL, ten autoimmune diseases - Behçet’s disease, coeliac disease, dermatitis herpetiformis, psoriasis, rheumatoid arthritis, sarcoidosis, systemic lupus erythematosus (SLE), Sjögren’s syndrome, systemic sclerosis, and type 1 diabetes (T1D) - were identified to be statistically significantly associated with an increased risk of NHL.^4^ Of these, coeliac disease, rheumatoid arthritis,

Sjögren’s syndrome, and SLE, were classified as presenting highly suggestive (i.e., *P* < 1×10^-6^, at least 1000 NHL cases, and largest study in the review reporting a nominally significant result) or convincing evidence (i.e., *P* < 1×10^-6^, at least 1000 NHL cases, largest study in the review reporting a nominally significant result, minimal between-study heterogeneity, and no evidence of publication bias) evidence for NHL risk.^4,5^ Autoimmune diseases have long been considered potential risk factors for NHL.^6,7^ Proposed mechanisms for the associations between autoimmune diseases and NHL include chronic inflammation, antigen stimulation, and overlapping genetic susceptibility.^8–11^ However, the associations identified by the umbrella review were from systematic reviews and meta-analyses with various study design and reporting limitations.

Furthermore, the systematic reviews and meta-analyses included only case-control and cohort studies, which are susceptible to multiple biases that limit their ability to evaluate causal relationships. For instance, confounding factors (e.g., socioeconomic status, family history of lymphoma, and infectious diseases) and reverse causation were often not considered by the previous studies.^12–18^ For some autoimmune diseases, such as sarcoidosis, it is often unclear if the autoimmune disease precedes or develops after NHL.^19–24^

One way of addressing the issue of residual confounding and reverse causation is through instrumental variable analyses with genetic instruments, often called Mendelian randomisation (MR) analyses.^25^ Because genetic variants are randomly assigned at conception and are not affected by external factors such as chronic diseases and lifestyle factors, MR analyses mimic randomized experiments and are less susceptible to confounding and reverse causation compared with conventional observational studies.^26,27^

Therefore, the aim of this study was to use the MR design to evaluate the associations between genetically predicted susceptibility to ten autoimmune diseases and the risk of NHL and NHL subtypes (follicular lymphoma, mature T/NK-cell lymphomas, non-follicular lymphoma, and other and unspecified types of NHL).^4^ Because nearly all observational studies on the associations between autoimmune diseases and NHL suggested a positive association,^4,9,12,13^ our hypothesis was that the genetically predicted susceptibility to each autoimmune disease was associated with an increased risk of NHL and at least one of the NHL subtypes.

## METHODS

This study is reported following the Strengthening the Reporting of Observational Studies in Epidemiology Using Mendelian Randomisation guidelines (STROBE-MR, **Supplement 1**).^28^ Although there is no pre-registered protocol for this study, the analyses were designed prior to the conduct of the study. Data were retrieved between June 2022 and June 2023, and analyses were conducted between June 2022 and October 2023. The manuscript was posted on *medRxiv*.

### Study design

Figure 1 shows a schematic summary of this two-sample MR study. Overall, we extracted summary level data from genome-wide association studies (GWASs) to find genetic instruments for our exposures of interest (i.e., exposure GWASs) and then investigated the associations of these genetic instruments with the outcomes of interest (i.e., outcome GWASs).^25^ For the genetic instruments to be valid, three core assumptions must be met:^29^ (1) the instruments are associated with the exposure of interest (the relevance assumption), (2) the instruments are not associated with any confounders of the exposure-outcome relationship (the independence assumption), and 3) the instruments are associated with the outcome only through the exposure (the exclusion restriction assumption).

The analyses were carried out in three steps: first, genetic instruments for each exposure of interest (i.e., autoimmune disease) were selected from relevant exposure GWASs. Second, for these genetic instruments, the genetic instrument-outcome associations were extracted from the relevant outcome GWASs. For each autoimmune disease, the exposure-outcome association (i.e., the Wald ratio) was estimated by dividing the genetic instrument-outcome association by the genetic instrument-exposure association. Finally, the Wald ratios were summarised using different techniques (See **Statistical analyses**). Due to data availability and in order to minimize confounding due to population stratification, we only considered subjects of European ancestry.^25^

### Instrumental variable selection for autoimmune diseases

To fulfill the relevance assumption, we used single nucleotide polymorphisms (SNPs) as genetic instruments and selected those from GWASs that were (1) strongly associated with the specific autoimmune disease at genome-wide significance (i.e., *P* < 5×10^-8^) and (2) independent of one-another (i.e., an R^2^ < 1×10^-3^). We used a less stringent criterion for significance level, *P* < 5×10^−6^ for dermatitis herpetiformis and Sjögren’s syndrome to ensure that we had at least five SNPs as genetic instruments for each autoimmune disease.

We evaluated the ten autoimmune diseases that were at least nominally significantly associated (i.e., *P* < 5×10^-2^) with risk of NHL according to a recent umbrella review:^4^ Behçet disease, coeliac disease, dermatitis herpetiformis, psoriasis, rheumatoid arthritis, sarcoidosis, Sjögren’s syndrome, SLE, systemic sclerosis, and T1D. These autoimmune diseases were all associated with an increased risk of NHL. Of these, coeliac disease, rheumatoid arthritis, Sjögren’s syndrome, and SLE, were classified as having highly suggestive (i.e., *P* < 1×10^-6^, at least 1000 NHL cases, and largest study in the review reporting a nominally significant result) or convincing evidence (i.e., *P* < 1×10^-6^, at least 1000 NHL cases, largest study in the review reporting a nominally significant result, minimal between-study heterogeneity, and no evidence of publication bias) in the umbrella review.^4^ For each autoimmune disease, we selected the GWAS that had the largest sample size. In situations where no distinct GWAS publication for an autoimmune disease was available, we used summary statistics from genome-wide association analyses from FinnGen Release 8 (https://r8.finngen.fi/, **Supplementary 2**).^30^ Study characteristics of all summary level GWAS datasets for the autoimmune diseases used in main analyses are shown in **Table 1**.

**Table 1.** GWAS datasets used for the autoimmune diseases in the in main analyses.

| Trait | Study | Countries | Cohorts | Number of cases | Number of controls | Number of SNPs | R<sup>2</sup> (% of variance explained) | Median *F* statistics (range) |
| --- | --- | --- | --- | --- | --- | --- | --- | --- |
| Behçet disease | Fernández et al 2021<sup>1</sup> | West Europe, Italy, Spain | Meta-analysis of three independent cohorts | 437 | 3,325 | 7 | 0.3 | 1 (0.1-7.9) |
| Coeliac disease | Dubois et al 2010<sup>2</sup> | The UK, the USA, Finland, Italy, Netherlands, Poland, Hungary, Spain | Meta-analysis of seven independent cohorts | 4,918 | 5,684 | 12 | 1.2 | 53 (30-96) |
| Dermatitis herpetiformis | Kurki et al 2023<sup>3</sup> | Finland | FinnGen | 435 | 341,188 | 8* | 1.3 | 22 (20-35) |
| Psoriasis | Tsoi et al 2017<sup>4</sup> | North America, Sweden | Meta-analysis of eight independent cohorts | 11,988 | 275,335 | 32 | 1.8 | 59 (30-310) |
| Rheumatoid arthritis | Ishigaki et al 2022<sup>5</sup> | The US, the UK, Canada, Spain, Sweden, the Netherlands, France | Meta-analysis of 25 independent cohorts | 22,350 | 74,823 | 59 | 2.4 | 42 (30-762) |
| Sarcoidosis | Kurki et al 2023<sup>3</sup> | Finland | FinnGen Study | 3,597 | 337,121 | 9 | 0.7 | 34 (32-116) |
| Sjögren's syndrome | Kurki et al 2023<sup>3</sup> | Finland | FinnGen Study | 2,735 | 332,115 | 11* | 0.8 | 23 (21-73) |
| Systemic lupus erythematosus | Wang et al 2021<sup>6</sup> | Spain, northern and western Europe, Italy | Meta-analysis of three independent cohorts | 4,576 | 8,039 | 24 | 2.5 | 44 (30-268) |
| Systemic sclerosis | López-Isac et al 2019<sup>7</sup> | Spain, Germany, the Netherlands, USA, France, Spain, Italy, UK, Sweden, Norway, Australia/UK | Meta-analysis of 14 independent cohorts | 9,095 | 17,584 | 21 | 1.3 | 41 (30-100) |
| Type I diabetes | Chiou et al 2021<sup>8</sup> | The USA, the UK, Ireland, Finland | Meta-analysis of nine independent cohorts including FinnGen Study | 18,942 | 501,638 | 72 | 3.1 | 45 (30-1077) |

Footnote: GWAS: genome-wide association study; ICD-10: International Classification of Diseases, Tenth Revision; NA: not available; the UK: the United Kingdoms; the USA: the United States.
\* $P < 5 \times 10^{-6}$ used as criteria to select genetic instruments.

Because human leukocyte antigen (HLA) region is highly predictive of both autoimmune diseases and NHL, we excluded SNPs from this locus from all analyses in order to prevent potential bias due to genetic pleiotropy.^31,32^ We defined the HLA region as location from 28,477,797 to 33,448,354 on chromosome 6 for Genome Reference Consortium Human Build 37.^33^

### Outcomes

Our primary outcome was risk of NHL. We selected the largest GWAS on NHL, which evaluated subjects from UK Biobank and Kaiser Permanente (**Table 2** and **Supplementary Text**).^34^ In secondary analyses for the genetically-predicted autoimmune diseases found to be nominally significantly associated with NHL (see **Statistical analyses**), we further evaluated the associations between these autoimmune diseases and NHL subtypes. We identified the NHL subtypes from FinnGen Release 8 and we selected four major NHL subtypes, consistent with the FinnGen classification system: follicular lymphoma, mature T/natural killer (NK)-cell lymphomas, non-follicular lymphoma, and other and unspecified types of NHL (**Table 2**).^30^ Of note, as the data on the NHL subtypes were extracted from a different dataset from the NHL GWAS, the secondary analyses also served as replication analyses using an independent dataset.^30^

**Table 2.** GWAS datasets used for the non-Hodgkin lymphoma in the in main analyses.

| **Trait** | **GWAS data source** | **Countries** | **Cohorts** | **Case definition** | **Control definition** | **Number of cases** | **Number of controls** |
| --- | --- | --- | --- | --- | --- | --- | --- |
| **Non-Hodgkin lymphoma** | Rashkin et al 2020<sup>9</sup> | The UK, the USA | UK Biobank, Kaiser Permanente cohorts | ICD-O-3 codes and SEER cite recode paradigm<sup>10</sup> | Individuals with no record of any cancer in the relevant registries | 2,400 | 410,350 |
| **Follicular lymphoma** | Kurki et al 2023<sup>3</sup> | Finland | FinnGen Study | ICD-10: C82 | Population controls | 955 | 271,463 |
| **Mature T/NK-cell lymphomas** | Kurki et al 2023<sup>3</sup> | Finland | FinnGen Study | ICD-10: C84 | Population controls | 296 | 271,463 |
| **Non-follicular lymphoma** | Kurki et al 2023<sup>3</sup> | Finland | FinnGen Study | ICD-10: C83 | Population controls | 2,340 | 271,463 |
| **Other and unspecified types of non-Hodgkin lymphoma** | Kurki et al 2023<sup>3</sup> | Finland | FinnGen Study | ICD-10: C85 | Population controls | 982 | 271,463 |

Footnote: GWAS: genome-wide association study; ICD-10: International Classification of Diseases, Tenth Revision; NA: not available; UK:
United Kingdoms; the USA: the United States.

### Statistical analyses

#### Main analyses and sensitivity analyses

For each autoimmune disease, we calculated the SNP-specific Wald ratio, defined as β_EXP-OUT_ = β_SNP-OUT_ / β_SNP-EXP_ (Figure 1). We used inverse-variance weighted (IVW) analysis as our main analyses to sum the Wald ratios, which assign weights to each SNP in inverse proportion to the variance of the β_SNP-OUT_, assuming all instruments to be valid.^35,36^ However, the IVW analysis may be biased if any of the included instruments are invalid (e.g., if the genetic instruments affect multiple traits, which is known as horizontal pleiotropy).^37^ Therefore, we carried out three sensitivity analyses that provide unbiased estimates even in the presence of some invalid instruments:^25,38^ MR-Egger regression,^36,39^ weighted mode estimator analysis,^40^ and weighted median estimator analysis (**Supplementary Text**).^41^ Furthermore, to address potential weak instrument bias, which may be introduced when the genetic variants explain a very small proportion of the variation in the exposure,^29^ we included robust adjusted profile scores (RAPS) as an additional sensitivity analysis (**Supplementary Text**).^25,42^

**Figure 1.**
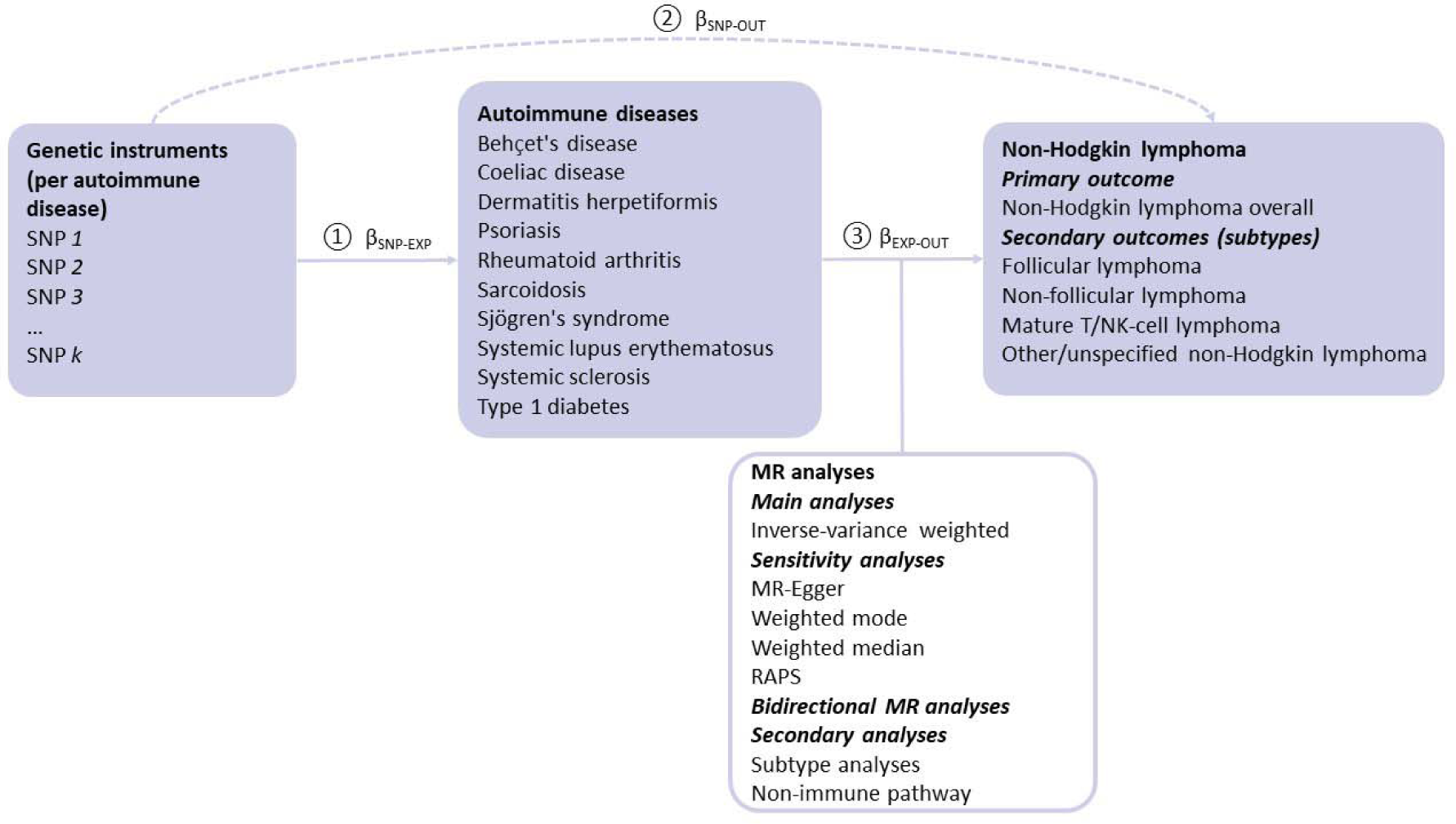
A schematic overview of the study design. Abbreviations: EXP: exposure; HLA: human leukocyte antigen; MR: mendelian randomisation; OUT: outcome; RAPS: robust adjusted profile scores; SNP: single nucleotide polymorphism. Wald ratio: 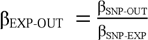; The analyses were carried out in three steps: Step 1, genetic instruments for each autoimmune disease were selected from relevant GWASs(i.e., *P* < 5 × 10^−8^ and an R^2^ <0.001) with β_SNP-EXP_ (i.e., association between genetic instrument and the exposure) extracted. Step 2, for these genetic instruments, the genetic instrument-outcome associations β_SNP-OUT_ were extracted from the outcome GWAS. Step 3, for each autoimmune disease, the Wald ratio (i.e., the causal estimate) was estimated for k number of SNPs by the formula β_EXP-OUT_ = β_SNP-OUT_/β_SNP-EXP_, and next summarised using the listed MR analyses. In the secondary analyses, only inverse-variance weighted method was applied.

We evaluated the strength of the instruments using R^2^ (i.e., proportion of variance of the exposure explained by the genetic instrument) and *F* statistics. In particular, we used the *get_r_from_bsen* function in the *TwoSampleMR* package in *R* (version 4.3.0), and summed the absolute values across the independent SNPs to estimate the composite R^2^ for each autoimmune disease. *F* statistics were calculated using the formula *F* statistic = 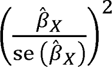, ^43^ where 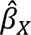 refers to the genetic association between the instrument X with the exposure, and se (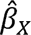) refers to the standard error of 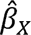.

#### Bidirectional analyses

To help establish the direction of effects between two traits,^25^ we carried out bidirectional analyses of genetically predicted NHL and risk of each autoimmune disease.^25^ Evidence of an effect in both directions could suggest that an effect acts in both directions between two traits so that changing one will change the other (i.e., the true bidirectional relationship). Otherwise it may suggest that a biasing pathway may be present.^25^ Evidence of an effect in one direction but not the other supports that a biasing pathway is less likely to be present.

#### Secondary analyses

To further validate the principal findings from the main analyses, we carried out two sets of secondary analyses for the autoimmune diseases that were nominally significantly associated with risk of NHL (i.e., T1D and sarcoidosis, see **Results**): (1) evaluating the risk of four NHL subtypes using a dataset independent from the main analyses and (2) restricting the genetic instruments to those that are less likely to affect immune function. The analyses of the NHL subtypes were carried out to identify which of the NHL subtypes were driving the observed association with NHL, and also to serve as a replication using a separate outcome study population not overlapping with the NHL study population of the other analyses. The analysis involving selected pathways were carried out to address horizontal pleiotropy. In particular, given the shared role of immune function in the pathway to developing both autoimmune diseases and NHL, we sought to identify biological pathways for the autoimmune diseases that were to a lesser degree linked to immune function (**Supplementary Table 1**). For T1D, we restricted this secondary analysis to SNPs linked to insulin production, while for sarcoidosis we selected SNPs that were not directly linked to immune function. The KEGG and GeneCards databases were used to identify the function of all T1D and sarcoidosis SNPs.^44,45^ For all secondary analyses, only IVW analyses were carried out.

#### Software

We used *TwoSampleMR* package in *R* (version 4.3.0) to run the two-sample MR analyses, and *forplo* to generate the forest plots.^46^ A *P* < 5×10^-2^ was considered nominally significant. To correct for multiple testing of the ten autoimmune diseases in the main analyses, the level for statistical significance was set at *P* < 5×10^−2^/10 = 5×10^−3^. To make the results more interpretable, all causal estimates were multiplied by 0.693 (= tay^2^) and next exponentiated in order to represent the odds ratios (ORs) for NHL per doubling in the prevalence of the autoimmune disease under study.^43^

#### Ethics

Only summary-level data from published studies with relevant ethical approvals were used in this study so approval from institutional review board was not necessary.

#### Patient and public involvement

No patients or members of the public were involved in the conception of the study, interpretation of the results, or drafting of the manuscript. We do not have plans to disseminate the results to research participants or relevant patient communities.

## RESULTS

The number of cases and controls identified in the relevant GWASs were 437 and 3,325 for Behçet’s disease, 4,918 and 5,684 for coeliac disease, 435 and 341,188 for dermatitis herpetiformis, 4,576 and 8,039 for lupus, 11,988 and 275,335 for psoriasis, 22,350 and 74,823 for rheumatoid arthritis, 3,597 and 337,121 for sarcoidosis, 2,735 and 332,115 for Sjögren’s syndrome, 9,095 and 17,584 for systemic sclerosis, 18,942 and 501,638 for type 1 diabetes, and 2,400 and 410,350 for NHL (**Table 1** and **Table 2**).^34^ The variance in the exposure explained by the genetic instruments ranged from 0.3% for Behçet disease to 3.1% for T1D (**Table 1**).

### Primary analyses

A doubling in the genetically-predicted prevalence of T1D was associated with an OR for NHL of 0.95 (95% confidence interval [CI]: 0.92 to 0.98, *P* = 5×10^-3^), while a doubling in the genetically-predicted prevalence of sarcoidosis was associated with an OR for NHL of 0.92 (95% CI: 0.85 to 0.99, *P* = 2.8×10^-2^) (Figure 2). We did not observe significant associations between the other eight autoimmune diseases and risk of NHL (Figure 2).

**Figure 2.**
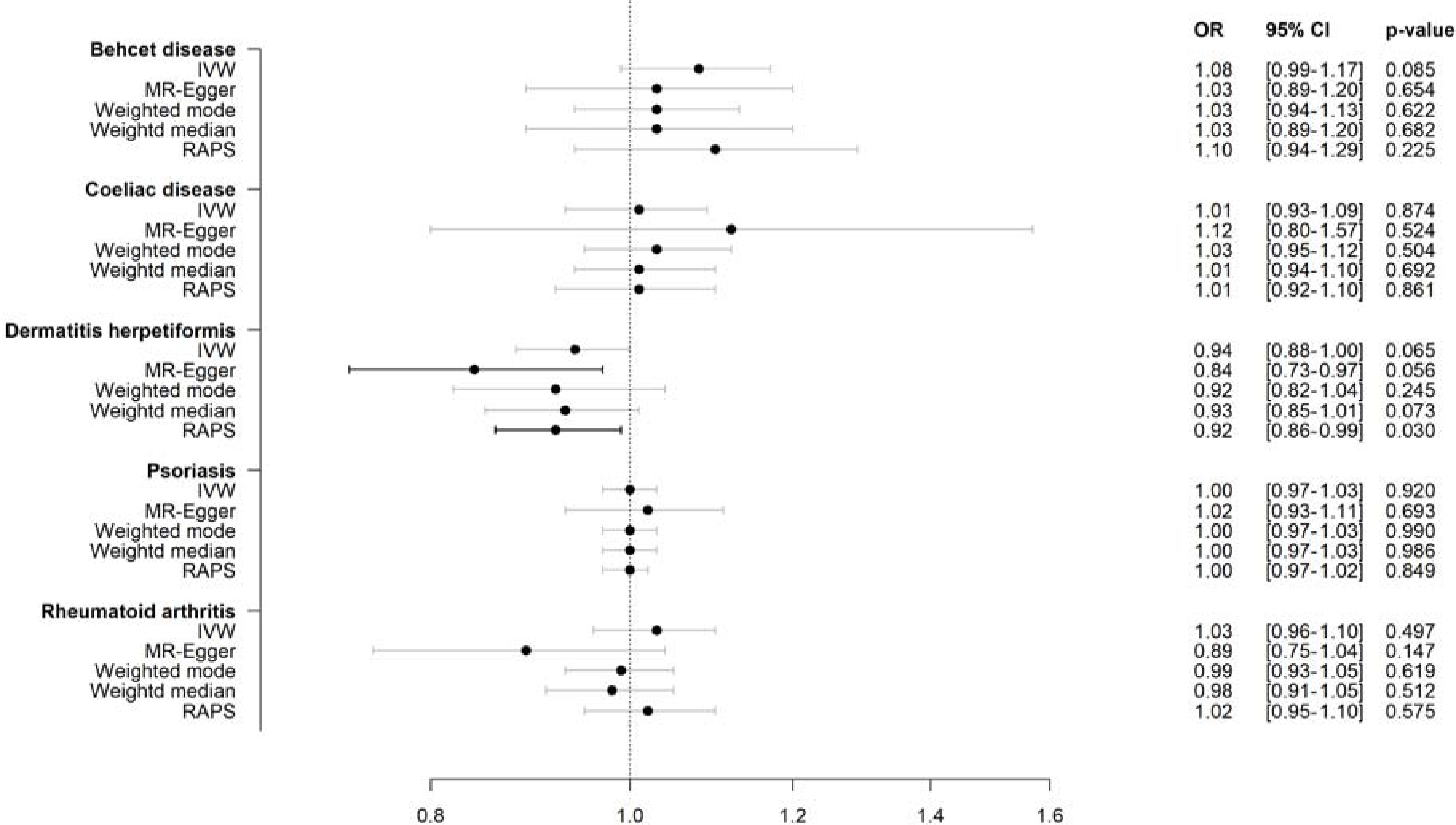
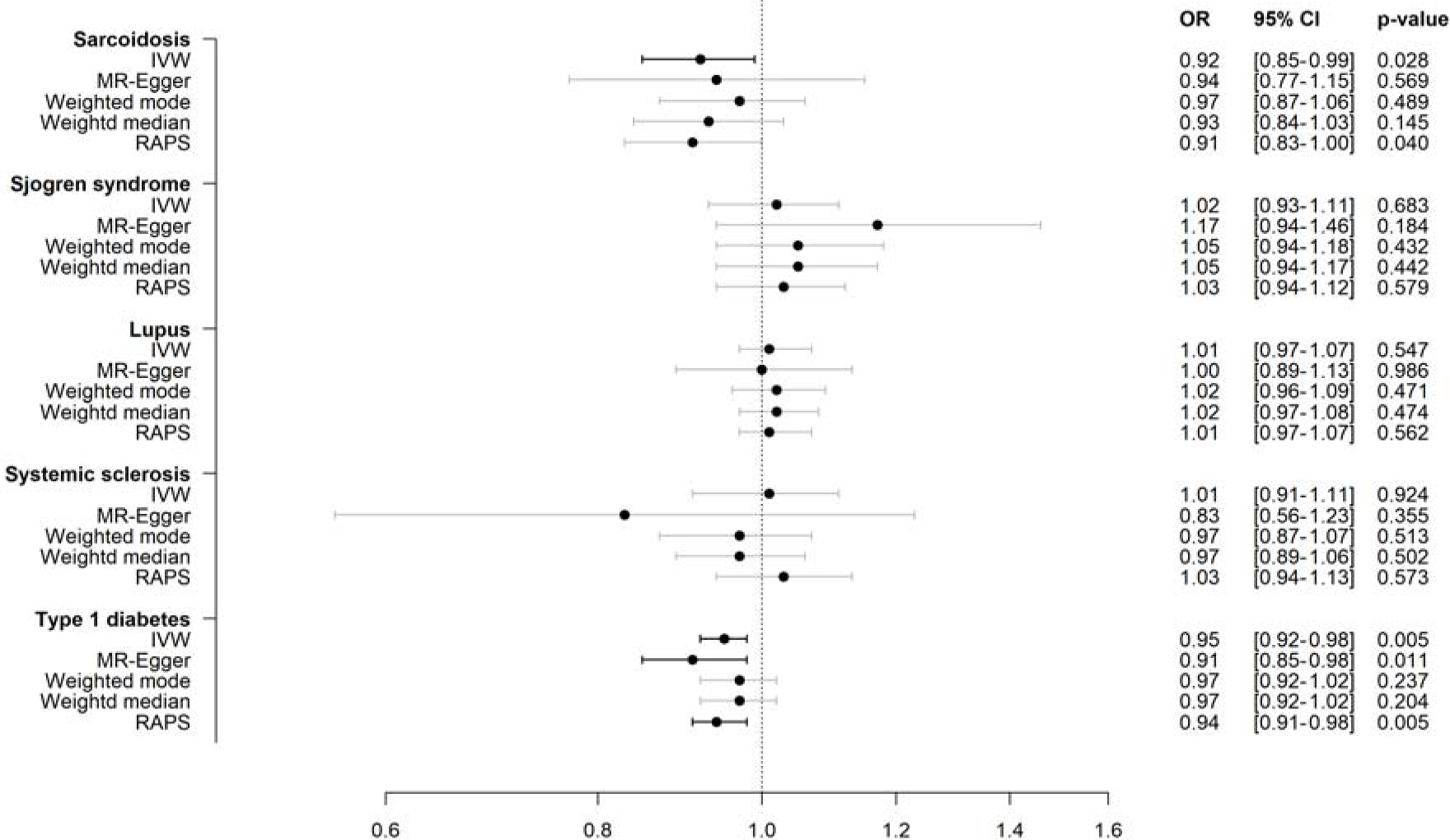
a. Genetically predicted susceptibility to ten autoimmune diseases and risk of non-Hodgkin lymphoma overall Abbreviations: CI: confidence interval; IVW: inverse-variance weighted; MR: mendelian randomisation; OR: odds ratio; RAPS: robust adjusted profile scores. Legend: all causal estimates were multiplied by 0.693 and next exponentiated in order to represent the odds ratios (ORs) for NHL per doubling in the prevalence of the autoimmune disease under study. b. Genetically predicted susceptibility to ten autoimmune diseases and risk of non-Hodgkin lymphoma overall Abbreviations: CI: confidence interval; IVW: inverse-variance weighted; MR: mendelian randomisation; OR: odds ratio; RAPS: robust adjusted profile scores. Legend: all causal estimates were multiplied by 0.693 and next exponentiated in order to represent the odds ratios (ORs) for NHL per doubling in the prevalence of the autoimmune disease under study.

### Sensitivity analyses

MR-Egger, weighted mode and weighted median yielded ORs comparable to those from the main analyses, and with overlapping CIs, indicating little presence of pleiotropy (Figure 2). Furthermore, the RAPS sensitivity analyses did not suggest bias due to weak instruments. For the bidirectional analyses we did not observe significant associations between NHL and the risk of any of the autoimmune diseases (**Supplementary Table 2**).

### Secondary analyses

In the secondary analyses, we evaluated the autoimmune diseases that were at least nominally significantly associated with risk of NHL, i.e., T1D and sarcoidosis. In the analyses restricted to non-immune pathways, we observed ORs of 0.96 (95% CI: 0.87 to 1.07) for T1D and 0.96 (95% CI: 0.88 to 1.06) for sarcoidosis, respectively, supporting the main analyses.

Figure 3 shows the IVW analyses of genetically predicted susceptibility to T1D and sarcoidosis, and the risk of NHL subtypes. For T1D, the association with composite NHL appeared to be driven by follicular lymphoma, with an OR of 0.91 (95% CI: 0.86 to 0.96, *P* = 1×10^-3^).

**Figure 3.**
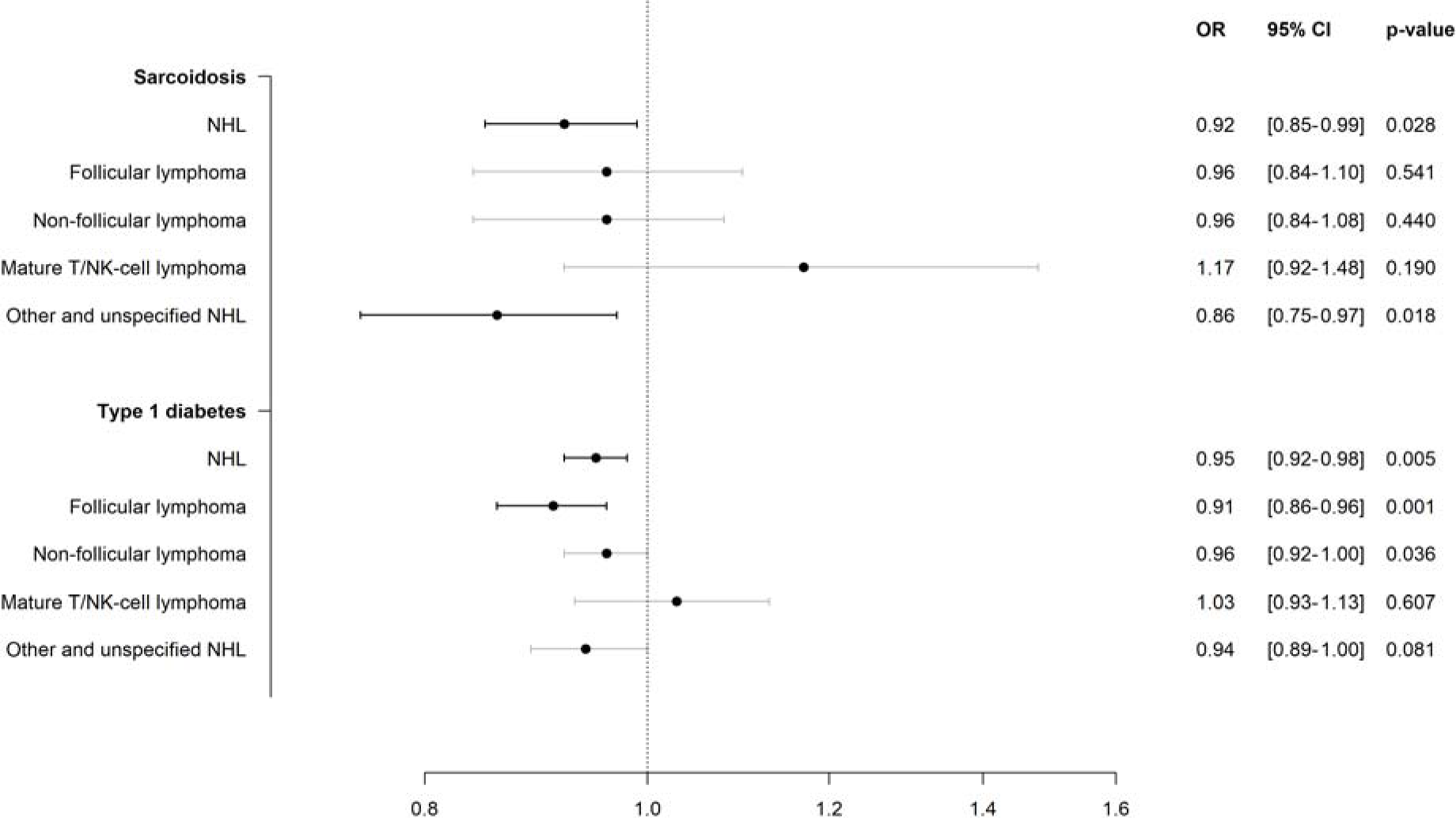
Inverse-variance weighted analyses of genetically predicted susceptibility to type 1 diabetes, and sarcoidosis, and risk of non-Hodgkin lymphoma subtypes. Abbreviations: CI: confidence interval; NHL: non-Hodgkin lymphoma; OR: odds ratio. Legend: 1. the analyses were based on inverse-variance weighted approach. 2. all causal estimates were multiplied by 0.693 and next exponentiated in order to represent the odds ratios (ORs) for NHL per doubling in the prevalence of the autoimmune disease under study. 3. As mentioned in Table 2, we used different datasets for NHL and NHL subtypes.

Sarcoidosis was most strongly associated with other and unspecified types of NHL, with an OR of 0.86 (95% CI: 0.75 to 0.97, *P* = 1.8×10^−2^).

## DISCUSSION

### Principal findings

In this MR study of ten autoimmune diseases previously linked to an increased risk of NHL, we found that genetically predicted susceptibility to T1D, and to some extent, genetically predicted susceptibility to sarcoidosis, were associated with a reduced risk of NHL. While these findings were consistent across a wide range of sensitivity analyses, no clear associations were observed between the other eight autoimmune diseases and risk of NHL. Using an approach that attempts to address potential residual confounding and reverse causation, our findings contradict those reported in previous traditional observational studies. This highlights the need for future studies with different datasets, approaches, and populations to further examine the potential associations between these autoimmune diseases and the risk of NHL.

### Context of primary findings

Autoimmune diseases have long been considered potential risk factors for NHL, especially rheumatoid arthritis, SLE, and Sjögren’s syndrome.^6,7,13,47,48^ A recent large-scale prospective cohort study found that among all cancers, lymphoma demonstrated the most extensive associations with different immune-mediated diseases.^9^ According to a previous umbrella review evaluating the associations between any environmental risk factors and the risk of NHL reported in published meta-analyses, there were consistent statistically significant associations between autoimmune diseases and NHL risk.^4^ Although the exact mechanisms for positive associations between autoimmune diseases and NHL remain unclear, there are several mechanisms proposed, including chronic inflammation, antigen stimulation, overlapping genetic susceptibility, and dysfunction of certain protein families.^8–11,49^ Furthermore, the observed associations between autoimmune diseases and NHL may be attributable to the immunosuppressants that are used as treatments as well.^47,50^

In our study, two autoimmune diseases -T1D and sarcoidosis - were found to be associated with NHL. However, contrary to our pre-specified hypotheses, which were based on the summary relative risks from an umbrella review (relative risk 1.55 (95% CI: 1.15 to 2.08) for associations between T1D and NHL, and relative risk 1.43 (95% CI: 1.03 to 1.99) for sarcoidosis and NHL),^4^ we found a statistically significant negative association between genetically predicted susceptibility to T1D and NHL. Although we also observed a negative association between genetically predicted susceptibility to sarcoidosis and NHL, the association was no longer significant after accounting for multiple testing. While no previous epidemiologic studies have reported statistically significant negative associations for either T1D or sarcoidosis with NHL,^8,9,13,22,51^ it has been suggested that the pathways involved in autoimmune disease and cancer development may work in opposite directions.^52^ Moreover, it remains unclear if sarcoidosis precedes or follows NHL, as several observational studies reported occurrence of sarcoidosis among NHL patients after treatment of NHL or concomitant occurrence of two diseases.^19–22^

There are several potential explanations for the discrepancy between our findings and those of the previous umbrella review. First, meta-analyses of observational studies and MR analyses have different bias structures. While traditional observational studies often suffer from confounding, misclassification, and selection bias, and meta-analyses are often susceptible to publication bias (i.e., the lack of publishing certain findings, often null findings), horizontal pleiotropy is of particular concern for MR studies. Second, many of the traditional observational studies and meta-analyses have relatively small sample sizes. In particular, the previous meta-analyses for T1D only included three individual observational studies with 1155 NHL cases and the meta-analysis for sarcoidosis only included seven studies with 150 sarcoidosis cases.^4,53^ Furthermore, according to a formal critical appraisal tool for systematic reviews and meta-analyses (i.e., AMSTAR 2), both meta-analyses were found to have at least one critical weakness.^4,5^ Third, given that genetic variants are fixed at conception, the exposure measured in MR analyses are typically interpreted as the lifelong exposure. In observational studies on the association between autoimmune diseases and NHL, the exposure is usually measured and interpreted in a defined time period from disease onset of autoimmune diseases. Finally, we conducted our study in population of European ancestry, while some meta-analyses investigated the associations in multiple populations other than European ancestry.^53,54^ However, the findings from populations with different ancestries did not differ drastically in the umbrella review.^53,54^

### Strengths and limitations of this study

The MR design is a major strength of our study, as it reduces the influence of residual confounding and reverse causation. In particular, confounding due to infectious diseases (an important risk factor for NHL and for autoimmune diseases) is minimized in this study. Moreover, to our knowledge, this is the first MR study that investigates the association between multiple autoimmune diseases and NHL.

Our study had several limitations. First, this study is limited to populations of European ancestry, which have the highest incidence of NHL, and it is unclear whether the findings can be generalised beyond this population. However, only GWAS data for populations of European ancestry are publicly available for all the autoimmune diseases, NHL, and NHL subtypes that we investigated.^3^ Furthermore, using the same population also ensures that the exposure dataset and outcome dataset are as similar as they could be. Second, NHL is a heterogenous group of haematologic disorders, with more than 20 subtypes.^50^ Although we used the most robust dataset for NHL subtypes (i.e., FinnGen), different categorisation of NHL subtypes is possible and may yield different results. Third, one autoimmune disease, Behçet disease, had suboptimal *F* statistics for its genetic instruments, which may cause weak instrument bias. However, when we conducted RAPS analysis to address potential bias due to weak instruments, the observed findings were similar to the main analyses.^25,42^ Lastly, given the shared genetic basis of autoimmune diseases and NHL,^11,55^ we were particularly cautious about potential bias due to horizontal pleiotropy by immune function. To minimize the impact of this bias, we excluded SNPs in the HLA region and carried out various sensitivity analyses. We also carried out secondary analyses for T1D and sarcoidosis restricted to the insulin and non-immune pathways, respectively. Although our sensitivity analyses did not suggest important bias due to genetic pleiotropy, it is still possible that potential pleiotropy may partially affect our study findings.

While our study addressed potential residual confounding that may have been present in previous observational studies, the MR design may be biased if the instrumental variable assumptions do not hold. Thus, the association between autoimmune diseases and risk of NHL needs further investigation with evidence triangulation using different datasets, populations, and approaches.

### Conclusions

This MR analysis found that genetically predicted susceptibility to T1D, and to some extent genetically predicted susceptibility to sarcoidosis, were associated with a lower risk of NHL. Future research using different datasets, approaches, and populations is necessary to develop a more comprehensive understanding of the associations between T1D and NHL, and sarcoidosis and NHL.

## Supporting information

Supplement 1_MR STROBE

## Data Availability

The data will be made available via a publicly accessible repository on publication.

## Contributors

XS, JDW and TR originally conceived this study. XS and TR designed this study. XS and TR acquired the data. XS and TR conducted the statistical analysis. XS drafted the manuscript. All authors participated in the interpretation of the data. All authors and critically revised the manuscript for important intellectual content. XS and TR had full access to all the data in the study and take responsibility for the integrity of the data and the accuracy of the data analysis. TR provided supervision. XS and TR are the guarantors. The corresponding author attests that all listed authors meet authorship criteria and that no others meeting the criteria have been omitted.

## Patient consent

Not required

## Ethical approval

Not required

## Data sharing

The data will be made available via a publicly accessible repository on publication. The code for analyses is included as supplementary information.

## Transparency

The senior author (manuscript guarantor) affirms that the manuscript is an honest, accurate, and transparent account of the study being reported; that no important aspects of the study have been omitted; and that any discrepancies from the study as planned (and, if relevant registered) have been explained.

## License

The Corresponding Author has the right to grant on behalf of all authors and does grant on behalf of all authors, a worldwide license to the Publishers and its licensees in perpetuity, in all forms, formats and median (whether known now or created in the future), to i) publish, reproduce, distribute, display and store the Contribution, ii) translate the Contribution into other languages, create adaptations, reprints, include within collections and create summaries, extracts and/or, abstracts of the Contribution, iii) create any other derivative work(s) based on the Contribution, iv) to exploit all subsidiary rights in the Contribution, v) the inclusion of electronic links from the Contribution to third party material where-ever it may be located; and, vi) license any third party to do any or all of the above. The default license, a CC BY NC license, is needed.

This is an Open Access article distributed in accordance with the Creative Commons Attribution Non Commercial (CC BY-NC 4.0) license, which permits others to distribute, remix, adapt, build upon this work non-commercially, and license their derivative works on different terms, provided the original work is properly cited and the use is non-commercial.

See: http://creativecommons.org/licenses/by-nc/4.0/.

## REFERENCES

1. Siegel RL, Miller KD, Wagle NS, Jemal A. Cancer statistics, 2023. CA Cancer J. Clin. 2023;73(1):17–48.

2. Mafra A, Laversanne M, Gospodarowicz M, et al. Global patterns of non-Hodgkin lymphoma in 2020. Int. J. Cancer. 2022;151(9):1474–1481.

3. Bispo JAB, Pinheiro PS, Kobetz EK. Epidemiology and Etiology of Leukemia and Lymphoma. Cold Spring Harb. Perspect. Med. 2019.

4. Shi X, Zhuo H, Du Y, Nyhan K, Ioannidis J, Wallach JD. Environmental risk factors for non-Hodgkin’s lymphoma: umbrella review and comparison of meta-analyses of summary and individual participant data. BMJ Medicine. 2022;1(1):e000184.

5. Shea BJ, Reeves BC, Wells G, et al. AMSTAR 2: a critical appraisal tool for systematic reviews that include randomised or non-randomised studies of healthcare interventions, or both. BMJ. 2017;358:j4008.

6. Zhang Y, Dai Y, Zheng T, Ma S. Risk Factors of Non-Hodgkin Lymphoma. Expert Opin. Med. Diagn. 2011;5(6):539–550.

7. Bowzyk Al-Naeeb A, Ajithkumar T, Behan S, Hodson DJ. Non-Hodgkin lymphoma. BMJ (Clinical research ed.). 2018;362:k3204–k3204.

8. Masetti R, Tiri A, Tignanelli A, et al. Autoimmunity and cancer. Autoimmun Rev. 2021;20(9):102882.

9. He MM, Lo CH, Wang K, et al. Immune-Mediated Diseases Associated With Cancer Risks. JAMA Oncol. 2022;8(2):209–219.

10. Baecklund E, Smedby KE, Sutton LA, Askling J, Rosenquist R. Lymphoma development in patients with autoimmune and inflammatory disorders--what are the driving forces? Semin. Cancer Biol. 2014;24:61–70.

11. Conde L, Bracci PM, Halperin E, Skibola CF. A search for overlapping genetic susceptibility loci between non-Hodgkin lymphoma and autoimmune diseases. Genomics. 2011;98(1):9–14.

12. Ekström Smedby K, Vajdic CM, Falster M, et al. Autoimmune disorders and risk of non-Hodgkin lymphoma subtypes: a pooled analysis within the InterLymph Consortium. Blood. 2008;111(8):4029–4038.

13. Fallah M, Liu X, Ji J, Försti A, Sundquist K, Hemminki K. Autoimmune diseases associated with non-Hodgkin lymphoma: a nationwide cohort study. Ann. Oncol. 2014;25(10):2025–2030.

14. Morton LM, Slager SL, Cerhan JR, et al. Etiologic heterogeneity among non-Hodgkin lymphoma subtypes: the InterLymph non-Hodgkin lymphoma subtypes project. Journal of the National Cancer Institute Monographs. 2014;2014(48):130–144.

15. Op de Beeck A, Eizirik DL. Viral infections in type 1 diabetes mellitus — why the β cells? Nature Reviews Endocrinology. 2016;12(5):263–273.

16. Chabay PA, Preciado MV. EBV primary infection in childhood and its relation to B-cell lymphoma development: a mini-review from a developing region. Int. J. Cancer. 2013;133(6):1286–1292.

17. Jeong SH. HBV infection as a risk factor for non-Hodgkin lymphoma. Lancet Oncol. 2010;11(9):806.

18. Getts DR, Chastain EM, Terry RL, Miller SD. Virus infection, antiviral immunity, and autoimmunity. Immunol. Rev. 2013;255(1):197–209.

19. London J, Grados A, Fermé C, et al. Sarcoidosis occurring after lymphoma: report of 14 patients and review of the literature. Medicine (Baltimore). 2014;93(21):e121.

20. Cho H, Yoon DH, Kim JH, et al. Occurrence of sarcoidosis after chemotherapy for non-Hodgkin lymphoma. Korean J. Intern. Med. 2016;31(3):605–607.

21. Chalayer É, Bachy E, Occelli P, et al. Sarcoidosis and lymphoma: a comparative study. QJM. 2015;108(11):871–878.

22. Bonifazi M, Renzoni EA, Lower EE. Sarcoidosis and maligancy: the chicken and the egg? Curr. Opin. Pulm. Med. 2021;27(5):455–462.

23. Hauswirth AW, Skrabs C, Schützinger C, Gaiger A, Lechner K, Jäger U. Autoimmune hemolytic anemias, Evans’ syndromes, and pure red cell aplasia in non-Hodgkin lymphomas. Leuk. Lymphoma. 2007;48(6):1139–1149.

24. Jardin F. Development of autoimmunity in lymphoma. Expert Rev. Clin. Immunol. 2008;4(2):247–266.

25. Sanderson E, Glymour MM, Holmes MV, et al. Mendelian randomization. Nature Reviews Methods Primers. 2022;2(1):1–21.

26. Haycock PC, Burgess S, Wade KH, Bowden J, Relton C, Davey Smith G. Best (but oft-forgotten) practices: the design, analysis, and interpretation of Mendelian randomization studies. Am. J. Clin. Nutr. 2016;103(4):965–978.

27. Smith GD, Ebrahim S. ’Mendelian randomization’: can genetic epidemiology contribute to understanding environmental determinants of disease? Int. J. Epidemiol. 2003;32(1):1–22.

28. Skrivankova VW, Richmond RC, Woolf BAR, et al. Strengthening the reporting of observational studies in epidemiology using mendelian randomisation (STROBE-MR): explanation and elaboration. BMJ. 2021;375:n2233.

29. Davies NM, Holmes MV, Davey Smith G. Reading Mendelian randomisation studies: a guide, glossary, and checklist for clinicians. BMJ. 2018;362:k601.

30. Kurki MI, Karjalainen J, Palta P, et al. FinnGen provides genetic insights from a well-phenotyped isolated population. Nature. 2023;613(7944):508–518.

31. Matzaraki V, Kumar V, Wijmenga C, Zhernakova A. The MHC locus and genetic susceptibility to autoimmune and infectious diseases. Genome Biol. 2017;18(1):1–21.

32. Zhong C, Cozen W, Bolanos R, Song J, Wang SS. The role of HLA variation in lymphoma aetiology and survival. J. Intern. Med. 2019;286(2):154–180.

33. Human Genome Region MHC. https://www.ncbi.nlm.nih.gov/grc/human/regions/MHC?asm=GRCh37. Accessed April 7, 2023.

34. Rashkin SR, Graff RE, Kachuri L, et al. Pan-cancer study detects genetic risk variants and shared genetic basis in two large cohorts. Nature Communications. 2020;11(1):4423.

35. Burgess S, Butterworth A, Thompson SG. Mendelian randomization analysis with multiple genetic variants using summarized data. Genet. Epidemiol. 2013;37(7):658–665.

36. Burgess S, Thompson SG. Interpreting findings from Mendelian randomization using the MR-Egger method. Eur. J. Epidemiol. 2017;32(5):377–389.

37. Hemani G, Bowden J, Davey Smith G. Evaluating the potential role of pleiotropy in Mendelian randomization studies. Hum. Mol. Genet. 2018;27(R2):R195–R208.

38. Burgess S, Davey Smith G, Davies NM, et al. Guidelines for performing Mendelian randomization investigations. Wellcome Open Res. 2019;4:186.

39. Bowden J, Davey Smith G, Burgess S. Mendelian randomization with invalid instruments: effect estimation and bias detection through Egger regression. Int. J. Epidemiol. 2015;44(2):512–525.

40. Hartwig FP, Davey Smith G, Bowden J. Robust inference in summary data Mendelian randomization via the zero modal pleiotropy assumption. Int. J. Epidemiol. 2017;46(6):1985–1998.

41. Bowden J, Davey Smith G, Haycock PC, Burgess S. Consistent estimation in Mendelian randomization with some invalid instruments using a weighted median estimator. Genet. Epidemiol. 2016;40(4):304–314.

42. Zhao Q, Wang J, Hemani G, Bowden J, Small DS. Statistical inference in two-sample summary-data Mendelian randomization using robust adjusted profile score. 2020.

43. Burgess S, Thompson SG. Mendelian randomization: methods for causal inference using genetic variants. CRC Press; 2021.

44. Kanehisa M, Furumichi M, Tanabe M, Sato Y, Morishima K. KEGG: new perspectives on genomes, pathways, diseases and drugs. Nucleic Acids Res. 2017;45(D1):D353–D361.

45. Stelzer G, Rosen N, Plaschkes I, et al. The GeneCards Suite: From Gene Data Mining to Disease Genome Sequence Analyses. Curr Protoc Bioinformatics. 2016;54:1 30 31–31 30 33.

46. Hemani G, Zheng J, Elsworth B, et al. The MR-Base platform supports systematic causal inference across the human phenome. eLife. 2018;7:e34408.

47. Zintzaras E, Voulgarelis M, Moutsopoulos HM. The risk of lymphoma development in autoimmune diseases: a meta-analysis. Arch. Intern. Med. 2005;165(20):2337–2344.

48. Cuttner J, Spiera H, Troy K, Wallenstein S. Autoimmune disease is a risk factor for the development of non-Hodgkin’s lymphoma. The Journal of Rheumatology. 2005;32(10):1884–1887.

49. Khanmohammadi S, Shabani M, Tabary M, Rayzan E, Rezaei N. Lymphoma in the setting of autoimmune diseases: A review of association and mechanisms. Crit. Rev. Oncol. Hematol. 2020;150:102945.

50. Armitage JO, Gascoyne RD, Lunning MA, Cavalli F. Non-Hodgkin lymphoma. Lancet.556 2017;390(10091):298–310.

51. Mellemkjaer L, Pfeiffer RM, Engels EA, et al. Autoimmune disease in individuals and close family members and susceptibility to non-Hodgkin’s lymphoma. Arthritis Rheum. 2008;58(3):657–666.

52. Sakowska J, Arcimowicz Ł, Jankowiak M, et al. Autoimmunity and Cancer—Two Sides of the Same Coin. Front. Immunol. 2022;13:793234.

53. Wang Y, Liu X, Yan P, Bi Y, Liu Y, Zhang ZJ. Association between type 1 and type 2 diabetes and risk of non-Hodgkin’s lymphoma: A meta-analysis of cohort studies. Diabetes Metab. 2020;46(1):8–19.

54. Simon TA, Thompson A, Gandhi KK, Hochberg MC, Suissa S. Incidence of malignancy in adult patients with rheumatoid arthritis: a meta-analysis. Arthritis Res. Ther. 2015;17(1):212.

55. Din L, Sheikh M, Kosaraju N, et al. Genetic overlap between autoimmune diseases and non-Hodgkin lymphoma subtypes. Genet. Epidemiol. 2019;43(7):844–863. 570

## References

1. Ortiz Fernández L, Coit P, Yilmaz V, et al. Genetic Association of a Gain-of-Function IFNGR1 Polymorphism and the Intergenic Region LNCAROD/DKK1 With Behçet’s Disease. Arthritis & Rheumatology. 2021;73(7):1244–1252.

2. Dubois PCA, Trynka G, Franke L, et al. Multiple common variants for celiac disease influencing immune gene expression. Nat. Genet. 2010;42(4):295–302.

3. Kurki MI, Karjalainen J, Palta P, et al. FinnGen provides genetic insights from a well-phenotyped isolated population. Nature. 2023;613(7944):508–518.

4. Tsoi LC, Stuart PE, Tian C, et al. Large scale meta-analysis characterizes genetic architecture for common psoriasis associated variants. Nature Communications. 2017;8(1):15382.

5. Ishigaki K, Sakaue S, Terao C, et al. Multi-ancestry genome-wide association analyses identify novel genetic mechanisms in rheumatoid arthritis. Nat. Genet. 2022;54(11):1640–1651.

6. Wang Y-F, Zhang Y, Lin Z, et al. Identification of 38 novel loci for systemic lupus erythematosus and genetic heterogeneity between ancestral groups. Nature Communications. 2021;12(1):772.

7. López-Isac E, Acosta-Herrera M, Kerick M, et al. GWAS for systemic sclerosis identifies multiple risk loci and highlights fibrotic and vasculopathy pathways. Nature Communications. 2019;10(1):4955.

8. Chiou J, Geusz RJ, Okino M-L, et al. Interpreting type 1 diabetes risk with genetics and single-cell epigenomics. Nature. 2021;594(7863):398–402.

9. Rashkin SR, Graff RE, Kachuri L, et al. Pan-cancer study detects genetic risk variants and shared genetic basis in two large cohorts. Nature Communications. 2020;11(1):4423.

10. SEER Site Recode. https://seer.cancer.gov/siterecode/icdo3_dwhoheme/index.html. Accessed April 6, 2023.

