## Supplement 2 for "Autoimmune diseases and risk of non-Hodgkin lymphoma: A Mendelian randomisation study"

Xiaoting Shi, PhD Candidate^1,4^; Joshua D. Wallach, Assistant Professor^1,2^; Xiaomei Ma, Professor^3^; Tormod Rogne, Assistant Professor^3,4^

^1^ Department of Environmental Health Sciences, Yale School of Public Health, New Haven, Connecticut, USA

^2^ Department of Epidemiology, Rollins School of Public Health, Emory University, Atlanta, Georgia, USA

^3^ Department of Chronic Diseases Epidemiology, Yale School of Public Health, New Haven, Connecticut, USA

^4^ Yale Center for Perinatal, Pediatric, and Environmental Epidemiology, Yale School of Public Health, New Haven, Connecticut, USA

Corresponding author:

Xiaoting Shi, MPhil

Department of Environmental Health Sciences, Yale School of Public Health, New Haven, Connecticut, USA

**Supplementary text**

***Included cohorts for non-Hodgkin lymphoma and its subtypes***

FinnGen is a project on genetic research launched in 2017, with the aim of collecting biological samples from 500,000 participants in Finland over six years. UK Biobank is a long-term biobank study lanuched in 2006 in the United Kingdom. It investigates how genetic and environmental exposures contribute to the development of disease. The Kaiser Permanente Genetic Epidemiology Research on Adult Health and Aging cohorts includes more than 100,000 individuals with genome-wide SNP data and a measure of telomere length that are linked to participants’ electronic health record data.

***Mendelian randomization (MR) methods***

Inverse-variance weighted (IVW) method assigns weights to each SNP in inverse proportion to the variance of the SNP-NHL effects, assuming all instruments to be valid. While IVW method is most powerful when all instruments are valid, MR-Egger is a statistical technique that can be used to test the violations of the assumptions and provide an unbiased effect estimate under a weaker set of assumptions. In particular, it allows all genetic instruments to have pleiotropic effects as long as the pleiotropic effects on risk of NHL are not correlated with the magnitude of the genetic instruments’ effects on autoimmune disease. Weighted mode/median estimator analyses are another widely-used sensitivity analyses in MR studies, where genetic instruments with more precise causal estimates are given more weight. When the majority of the instruments do not have pleiotropic effects, weighted mode/median estimator analysis may be able to provide more reliable estimates than MR-Egger analysis. Robust adjusted profile scores (RAPS) is an MR method that maximises the profile likelihood of the Wald ratio, accounting for weak instrument bias, pleiotropy and extreme outliers.

| **Supplementary Table 1. SNPs used for nonimmune function analyses** | | | |
| --- | --- | --- | --- |
| **Autoimmune diseases** | **SNPs** | **Gene** | **Function in GeneCards*** |
| Sarcoidosis | rs1002015 | ACOXL | Acyl-CoA Oxidase Like |
| Sarcoidosis | rs1079242 | ANXA11 | Annexin A11 |
| Sarcoidosis | rs1860217 | GATSL3 | Cytosolic Arginine Sensor For MTORC1 Subunit 1 |
| Sarcoidosis | rs7581601 | FAM117B | Family With Sequence Similarity 117 Member B |
| Type 1 diabetes | rs55893453 | ADCY3 | Adenylate Cyclase 3 |
| Type 1 diabetes | rs7110099 | INS-IGF2 | Insulin- insulin Like Growth Factor 2 |
| Type 1 diabetes | rs9394158 | ITPR3 | Inositol 1,4,5-Trisphosphate Receptor Type 3 |
| Abbreviations: SNP: Single nucleotide polymorphism. | | | |
| Notes: *https://www.genecards.org/ | | | |

**Supplementary Table 2. Bidirectional analyses excluding HLA**

| **Autoimmune diseases** | **Analytical methods** | **Number of SNPs** | **Data source** | **ORs (95% CIs)*** | **P-value** |
| --- | --- | --- | --- | --- | --- |
| **Behcet's disease** | Inverse variance weighted | / | / | / | / |
|  | Mendelian randomization-Egger | / | / | / | / |
|  | Weighted mode | / | / | / | / |
|  | Weighted median | / | / | / | / |
|  | RAPS | / | / | / | / |
| **Coeliac disease** | Inverse variance weighted | 19 | FinnGen | 1.01 (0.92, 1.11) | 0.878 |
|  | Mendelian randomization-Egger | 19 | FinnGen | 1.11 (0.89, 1.39) | 0.347 |
|  | Weighted mode | 19 | FinnGen | 1.02 (0.89, 1.16) | 0.774 |
|  | Weighted median | 19 | FinnGen | 1.01 (0.92, 1.13) | 0.782 |
|  | RAPS | 19 | FinnGen | 1.04 (0.92, 1.17) | 0.534 |
| **Dermatitis herpetiformis** | Inverse variance weighted | 19 | FinnGen | 1.08 (0.91, 1.29) | 0.393 |
|  | Mendelian randomization-Egger | 19 | FinnGen | 1.21 (0.80, 1.85) | 0.389 |
|  | Weighted mode | 19 | FinnGen | 0.99 (0.69, 1.41) | 0.944 |
|  | Weighted median | 19 | FinnGen | 1.02 (0.80, 1.32) | 0.859 |
|  | RAPS | 19 | FinnGen | 1.09 (0.78, 1.50) | 0.622 |
| **Psoriasis** | Inverse variance weighted | 19 | FinnGen | 0.98 (0.93, 1.03) | 0.334 |
|  | Mendelian randomization-Egger | 19 | FinnGen | 0.95 (0.84, 1.08) | 0.428 |
|  | Weighted mode | 19 | FinnGen | 1.00 (0.91, 1.10) | 0.931 |
|  | Weighted median | 19 | FinnGen | 0.98 (0.93, 1.05) | 0.620 |
|  | RAPS | 19 | FinnGen | 0.96 (0.90, 1.02) | 0.216 |
| **Rheumatoid arthritis** | Inverse variance weighted | 19 | Ishigaki et al 2022 | 0.99 (0.89, 1.09) | 0.783 |
|  | Mendelian randomization-Egger | 19 | Ishigaki et al 2022 | 0.98 (0.75, 1.29) | 0.889 |
|  | Weighted mode | 19 | Ishigaki et al 2022 | 1.01 (0.92, 1.12) | 0.738 |
|  | Weighted median | 19 | Ishigaki et al 2022 | 0.99 (0.93, 1.07) | 0.921 |
|  | RAPS | 19 | Ishigaki et al 2022 | 1.02 (0.93, 1.12) | 0.692 |
| **Sarcoidosis** | Inverse variance weighted | 19 | FinnGen | 0.99 (0.94, 1.06) | 0.886 |
|  | Mendelian randomization-Egger | 19 | FinnGen | 0.95 (0.82, 1.11) | 0.527 |
|  | Weighted mode | 19 | FinnGen | 1.08 (0.94, 1.23) | 0.326 |
|  | Weighted median | 19 | FinnGen | 1.06 (0.97, 1.15) | 0.181 |
|  | RAPS | 19 | FinnGen | 1.00 (0.90, 1.10) | 0.956 |
| **Sjogren's syndrome** | Inverse variance weighted | 19 | FinnGen | 1.01 (0.94, 1.09) | 0.844 |
|  | Mendelian randomization-Egger | 19 | FinnGen | 1.10 (0.92, 1.32) | 0.308 |
|  | Weighted mode | 19 | FinnGen | 1.10 (0.92, 1.31) | 0.327 |
|  | Weighted median | 19 | FinnGen | 1.02 (0.92, 1.13) | 0.714 |
|  | RAPS | 19 | FinnGen | 1.01 (0.89, 1.14) | 0.849 |
| **Systemic lupus erythematosus** | Inverse variance weighted | 19 | FinnGen | 1.02 (0.91, 1.15) | 0.726 |
|  | Mendelian randomization-Egger | 19 | FinnGen | 1.12 (0.85, 1.49) | 0.432 |
|  | Weighted mode | 19 | FinnGen | 1.00 (0.79, 1.26) | 0.977 |
|  | Weighted median | 19 | FinnGen | 1.02 (0.86, 1.20) | 0.851 |
|  | RAPS | 19 | FinnGen | 1.03 (0.85, 1.25) | 0.779 |
| **Systemic sclerosis** | Inverse variance weighted | 19 | FinnGen | 1.09 (0.93, 1.28) | 0.277 |
|  | Mendelian randomization-Egger | 19 | FinnGen | 0.87 (0.60, 1.26) | 0.475 |
|  | Weighted mode | 19 | FinnGen | 1.04 (0.69, 1.57) | 0.853 |
|  | Weighted median | 19 | FinnGen | 1.03 (0.82, 1.29) | 0.801 |
|  | RAPS | 19 | FinnGen | 1.16 (0.87, 1.53) | 0.313 |
| **Type 1 diabetes** | Inverse variance weighted | 21 | Chiou et al 2021 | 0.99 (0.93, 1.04) | 0.568 |
|  | Mendelian randomization-Egger | 21 | Chiou et al 2021 | 1.13 (1.02, 1.25) | 0.031 |
|  | Weighted mode | 21 | Chiou et al 2021 | 1.04 (0.96, 1.13) | 0.327 |
|  | Weighted median | 21 | Chiou et al 2021 | 1.03 (0.98, 1.10) | 0.210 |
|  | RAPS | 21 | Chiou et al 2021 | 1.03 (0.97, 1.10) | 0.299 |
| Abbreviations: CI: confidence interval; HLA: human leukocyte antigen; OR: odds ratio; RAPS: Robust adjusted profile score; SNP: Single-nucleotide polymorphism. | | | | | |
| Notes: 1. We used a less stringent criterion for significance level (i.e., P value<5 ×10-6) in bidirectional analyses to ensure a decent amount of SNPs are identified as instruments. | | | | | |
| 2. We prioritized using the same data source from main analyses in bidirectional analyses. In case of insufficient information (e.g., missing non effect alleles), GWAS from FinnGen was used. We were not able to run bidirectional analyses for Behcet's disease due to the lack of information in the Fernández et al 2021 study, and no eligible GWAS from FinnGen. | | | | | |
| 3. * per doubling prevalence of the autoimmune disease | | | | | |

**Example code for the Mendelian randomisation analysis between type 1 diabetes and non-Hodgkin lymphoma**

library(TwoSampleMR)

library(mr.raps)

library(dplyr)

library(tidyverse)

\# LOAD EXPOSURE DATA

t1d_exp_dat <- read_exposure_data(

filename = "GCST90014023_buildGRCh38_5e8.txt",

sep = "\t",

snp_col = "variant_id",

beta_col = "beta",

effect_allele_col = "effect_allele",

other_allele_col = "other_allele",

eaf_col = "effect_allele_frequency",

pval_col = "p_value",

se_col = "standard_error"

)

t1d_exp_dat$exposure <- "T1D"

\# STEP CLUMPING

t1d_exp_dat<- clump_data(t1d_exp_dat)

\# LOAD OUTCOME DATA for NHL

nhl_out_dat <- read_outcome_data(

snps = t1d_exp_dat$SNP,

filename = "nonHogdkin_Rashkin2020_wBetaSE.txt",

sep = "\t",

snp_col = "variant_id",

beta_col = "Beta",

se_col = "SE",

effect_allele_col = "effect_allele",

other_allele_col = "other_allele",

pval_col = "p_value",

chr_col = "chromosome",

pos_col = "base_pair_location"

)

nhl_out_dat$outcome <- "NHL"

\# STEP HARMONIZE DATA

dat_t1d <- harmonise_data(

exposure_dat = t1d_exp_dat,

outcome_dat = nhl_out_dat

)

\# Manually INCLUDE SNP(s) THAT WERE EXCLUDED FOR BEING PALINDROMIC

dat_t1d$mr_keep[dat_t1d$SNP==c("rs112647257")|dat_t1d$SNP==c("rs12644686")|dat_t1d$SNP==c("rs17106304")|dat_t1d$SNP==c("rs17323934")|dat_t1d$SNP==c("rs1881146")|dat_t1d$SNP==c("rs2303137")|dat_t1d$SNP==c("rs34536443")|dat_t1d$SNP==c("rs41295159")|dat_t1d$SNP==c("rs4490209")|dat_t1d$SNP==c("rs55993634")|dat_t1d$SNP==c("rs6931296")|dat_t1d$SNP==c("rs78325861")|dat_t1d$SNP==c("rs7936434")|dat_t1d$SNP==c("rs8046043")] <- TRUE

\# EXCLUDE SNPs AT HLA REGION

dat_t1d$mr_keep[dat_t1d$SNP==c("rs1008438")|dat_t1d$SNP==c("rs112647257")|dat_t1d$SNP==c("rs2395471")|dat_t1d$SNP==c("rs2523679")|dat_t1d$SNP==c("rs6931296")|dat_t1d$SNP==c("rs72838204")] <- FALSE

\# STEP SELECT INSTRUMENTS

dat_t1d$mr_keep[dat_t1d$pval.exposure>5e-8] <- FALSE

\# MAIN ANALYSES

res_single <- mr_singlesnp(dat_t1d, all_method = c("mr_ivw"))

res <- mr(dat_t1d, method_list=c("mr_ivw"))

generate_odds_ratios(res)

\# SENSITIVITY ANALYSES

res2<-mr(dat_t1d, method_list=c("mr_ivw", "mr_egger_regression", "mr_weighted_mode","mr_weighted_median","mr_raps"))

generate_odds_ratios(res2)
