## Supplement 1_MR STROBE for "Autoimmune diseases and risk of non-Hodgkin lymphoma: A Mendelian randomisation study"

**STROBE-MR checklist of recommended items to address in reports of Mendelian randomization studies**^1^ ^2^

| **Item No.** | **Section** | **Checklist item** | **Page No.** | **Relevant text from manuscript** |
| --- | --- | --- | --- | --- |
| 1 | **TITLE and ABSTRACT** | Indicate Mendelian randomization (MR) as the study’s design in the title and/or the abstract if that is a main purpose of the study | 1 and 3 | Autoimmune diseases and risk of non-Hodgkin lymphoma: A Mendelian randomisation study |
|  | **INTRODUCTION** |  |  |  |
| 2 | **Background** | Explain the scientific background and rationale for the reported study. What is the exposure? Is a potential causal relationship between exposure and outcome plausible? Justify why MR is a helpful method to address the study question | 6-7 | “One way of addressing the issue of residual confounding and reverse causation is through instrumental variable analyses with genetic instruments, often called Mendelian randomisation (MR) analyses. Because genetic variants are randomly assigned at conception and are not affected by external factors such as chronic diseases and lifestyle factors, MR analyses mimic randomized experiments and are less susceptible to confounding and reverse causation compared with conventional observational studies.” |
| 3 | **Objectives** | State specific objectives clearly, including pre-specified causal hypotheses (if any). State that MR is a method that, under specific assumptions, intends to estimate causal effects | 7 | “Therefore, the aim of this study was to use the MR design to evaluate the associations between genetically-predicted susceptibility to ten autoimmune diseases and the risk of NHL and NHL subtypes (follicular lymphoma, mature T/NK-cell lymphomas, non-follicular lymphoma, and other and unspecified types of NHL). Because nearly all observational studies on the associations between autoimmune diseases and NHL suggested a positive association, our hypothesis was that the genetically-predicted susceptibility to each autoimmune disease was associated with an increased risk of NHL and at least one of the NHL subtypes.” |
|  | **METHODS** |  |  |  |
| 4 | **Study design and data sources** | Present key elements of the study design early in the article. Consider including a table listing sources of data for all phases of the study. For each data source contributing to the analysis, describe the following: |  |  |
|  | a) | Setting: Describe the study design and the underlying population, if possible. Describe the setting, locations, and relevant dates, including periods of recruitment, exposure, follow-up, and data collection, when available. | 8-10 | “Data were retrieved between June 2022 and June 2023, and analyses were conducted between June 2022 and October 2023”  …”We evaluated the ten autoimmune diseases that were at least nominally significantly associated (i.e., P < 5×10-2) with risk of NHL according to a recent umbrella review:4 Behçet disease, coeliac disease, dermatitis herpetiformis, psoriasis, rheumatoid arthritis, sarcoidosis, Sjögren's syndrome, SLE, systemic sclerosis, and T1D”…  …”We identified the NHL subtypes from FinnGen Release 8 and we selected four major NHL subtypes, consistent with the FinnGen classification system: follicular lymphoma, mature T/natural killer (NK)-cell lymphomas, non-follicular lymphoma, and other and unspecified types of NHL (Table 2)”…  See also table 1 and table 2. |
|  | b) | Participants: Give the eligibility criteria, and the sources and methods of selection of participants. Report the sample size, and whether any power or sample size calculations were carried out prior to the main analysis | 8-10 | See table 1 and table 2  We did not conduct power or sample size calculations prior to the main analysis. |
|  | c) | Describe measurement, quality control and selection of genetic variants | 9-10 | “To fulfill the relevance assumption, we used single nucleotide polymorphisms (SNPs) as genetic instruments and selected those from GWASs that were 1) strongly associated with the specific autoimmune disease at genome-wide significance (i.e., P < 5 ×10-8) and (2) independent of one-another (i.e., an R2 < 1 ×10-3). We used a less stringent criterion for significance level, P < 5 ×10-6 for dermatitis herpetiformis and Sjögren’s syndrome to ensure that we had at least five SNPs as genetic instruments for each autoimmune disease.” |
|  | d) | For each exposure, outcome, and other relevant variables, describe methods of assessment and diagnostic criteria for diseases | 8-10 | See table 1 and table 2 |
|  | e) | Provide details of ethics committee approval and participant informed consent, if relevant | 13 | “Ethics  Only summary-level data from published studies with relevant ethical approvals were used in this study so approval from institutional review board was not necessary.  Patient and public involvement  No patients or members of the public were involved in the conception of the study, interpretation of the results, or drafting of the manuscript. We do not have plans to disseminate the results to research participants or relevant patient communities.” |
| 5 | **Assumptions** | Explicitly state the three core IV assumptions for the main analysis (relevance, independence and exclusion restriction) as well assumptions for any additional or sensitivity analysis | 8 | “For the genetic instruments to be valid, three core assumptions must be met:29 1) the instruments are associated with the exposure of interest (the relevance assumption), 2) the instruments are not associated with any confounders of the exposure-outcome relationship (the independence assumption), and 3) the instruments are associated with the outcome only through the exposure (the exclusion restriction assumption). ” |
| 6 | **Statistical methods: main analysis** | Describe statistical methods and statistics used |  |  |
|  | a) | Describe how quantitative variables were handled in the analyses (i.e., scale, units, model) | 13 | “To make the results more interpretable, all causal estimates were multiplied by 0.693 (= log e2) and next exponentiated in order to represent the odds ratios (ORs) for NHL per doubling in the prevalence of the autoimmune disease under study.” |
|  | b) | Describe how genetic variants were handled in the analyses and, if applicable, how their weights were selected | 11 | “We used inverse-variance weighted (IVW) analysis as our main analyses to sum the Wald ratios, which assign weights to each SNP in inverse proportion to the variance of the βSNP-OUT, assuming all instruments to be valid.” |
|  | c) | Describe the MR estimator (e.g. two-stage least squares, Wald ratio) and related statistics. Detail the included covariates and, in case of two-sample MR, whether the same covariate set was used for adjustment in the two samples | 8-12 | “For each autoimmune disease, the exposure-outcome association (i.e., the Wald ratio) was estimated by dividing the genetic instrument-outcome association by the genetic instrument-exposure association. Finally, the Wald ratios were summarised using different techniques (See Statistical analyses).”  …”For each autoimmune disease, we calculated the SNP-specific Wald ratio, defined as βEXP-OUT = βSNP-OUT / βSNP-EXP (Figure 1). We used inverse-variance weighted (IVW) analysis as our main analyses to sum the Wald ratios, which assign weights to each SNP in inverse proportion to the variance of the βSNP-OUT, assuming all instruments to be valid” |
|  | d) | Explain how missing data were addressed | 8-12 | Missing effect or non effect alleles are inferred. |
|  | e) | If applicable, indicate how multiple testing was addressed | 13 | “To correct for multiple testing of the ten autoimmune diseases in the main analyses, the level for statistical significance was set at P < 5×10-2/10 = 5×10-3.” |
| 7 | **Assessment of assumptions** | Describe any methods or prior knowledge used to assess the assumptions or justify their validity | 11-12 | “We evaluated the strength of the instruments using R2 (i.e., proportion of variance of the exposure explained by the genetic instrument) and F statistics. In particular, we used the get_r_from_bsen function in the TwoSampleMR package in R (version 4.3.0), and summed the absolute values across the independent SNPs to estimate the composite R2 for each autoimmune disease. F statistics were calculated using the formula F statistic$=\left( \frac{\hat{\beta}_{X}}{se\left( \hat{\beta}_{X} \right)} \right)^{2}$,43 where $\hat{\beta}_{X}$refers to the genetic association between the instrument X with the exposure, and se$\hat{(\beta}_{X)}$ refers to the standard error of $\hat{\beta}_{X}$.” |
| 8 | **Sensitivity analyses and additional analyses** | Describe any sensitivity analyses or additional analyses performed (e.g. comparison of effect estimates from different approaches, independent replication, bias analytic techniques, validation of instruments, simulations) | 11-12 | “We used inverse-variance weighted (IVW) analysis as our main analyses to sum the Wald ratios, which assign weights to each SNP in inverse proportion to the variance of the βSNP-OUT, assuming all instruments to be valid.35,36 However, the IVW analysis may be biased if any of the included instruments are invalid (e.g., if the genetic instruments affect multiple traits, which is known as horizontal pleiotropy).37 Therefore, we carried out three sensitivity analyses that provide unbiased estimates even in the presence of some invalid instruments:25,38 MR-Egger regression,36,39 weighted mode estimator analysis,40 and weighted median estimator analysis (Supplementary Text).41 Furthermore, to address potential weak instrument bias, which may be introduced when the genetic variants explain a very small proportion of the variation in the exposure,29 we included robust adjusted profile scores (RAPS) as an additional sensitivity analysis (Supplementary Text).25,42” |
| 9 | **Software and pre-registration** |  |  |  |
|  | a) | Name statistical software and package(s), including version and settings used | 11 | …”we used the get_r_from_bsen function in the TwoSampleMR package in R (version 4.3.0)”… |
|  | b) | State whether the study protocol and details were pre-registered (as well as when and where) | 8 | “Although there is no pre-registered protocol for this study, the analyses were designed prior to the conduct of the study. Data were retrieved between June 2022 and June 2023, and analyses were conducted between June 2022 and October 2023. The manuscript was posted on medRxiv” |
|  | **RESULTS** |  |  |  |
| 10 | **Descriptive data** |  |  |  |
|  | a) | Report the numbers of individuals at each stage of included studies and reasons for exclusion. Consider use of a flow diagram | Not applicable |  |
|  | b) | Report summary statistics for phenotypic exposure(s), outcome(s), and other relevant variables (e.g. means, SDs, proportions) |  | See table 1 and table 2 |
|  | c) | If the data sources include meta-analyses of previous studies, provide the assessments of heterogeneity across these studies | Not applicable |  |
|  | d) | For two-sample MR:  i.  Provide justification of the similarity of the genetic variant-exposure associations between the exposure and outcome samples  ii.  Provide information on the number of individuals who overlap between the exposure and outcome studies |  | Due to the lack of information on effect allele frequencies in the outcome (NHL) sample, we were unable to compare the effect allele frequencies similarity of the genetic variant-exposure associations between the exposure and outcome samples. Nevertheless, since the participants from exposure and outcome samples are all both European ancestry, we would expect similar SNP-exposure associations in all samples. |
| 11 | **Main results** |  |  |  |
|  | a) | Report the associations between genetic variant and exposure, and between genetic variant and outcome, preferably on an interpretable scale | 14 | “The variance in the exposure explained by the genetic instruments ranged from 0.3% for Behçet disease to 3.1% for T1D (Table 1).”  See table 1. |
|  | b) | Report MR estimates of the relationship between exposure and outcome, and the measures of uncertainty from the MR analysis, on an interpretable scale, such as odds ratio or relative risk per SD difference | 14 | “A doubling in the genetically-predicted prevalence of T1D was associated with an OR for NHL of 0.95 (95% confidence interval [CI]: 0.92 to 0.98, P = 5×10-3), while a doubling in the genetically-predicted prevalence of sarcoidosis was associated with an OR for NHL of 0.92 (95% CI: 0.85 to 0.99, P = 2.8×10-2) (Figure 2). We did not observe significant associations between the other eight autoimmune diseases and risk of NHL (Figure 2).” |
|  | c) | If relevant, consider translating estimates of relative risk into absolute risk for a meaningful time period | Not applicable |  |
|  | d) | Consider plots to visualize results (e.g. forest plot, scatterplot of associations between genetic variants and outcome versus between genetic variants and exposure) |  | See Figure 2 and figure 3 |
| 12 | **Assessment of assumptions** |  |  |  |
|  | a) | Report the assessment of the validity of the assumptions | 14 | “MR-Egger, weighted mode and weighted median yielded ORs comparable to those from the main analyses, and with overlapping CIs, indicating little presence of pleiotropy (Figure 2). Furthermore, the RAPS sensitivity analyses did not suggest bias due to weak instruments. For the bidirectional analyses we did not observe significant associations between NHL and the risk of any of the autoimmune diseases (Supplementary Table 2).”  See Figure 2 |
|  | b) | Report any additional statistics (e.g., assessments of heterogeneity across genetic variants, such as *I^2^*, Q statistic or E-value) | Not applicable |  |
| 13 | **Sensitivity analyses and additional analyses** |  |  |  |
|  | a) | Report any sensitivity analyses to assess the robustness of the main results to violations of the assumptions | 14 | “MR-Egger, weighted mode and weighted median yielded ORs comparable to those from the main analyses, and with overlapping CIs, indicating little presence of pleiotropy (Figure 2). Furthermore, the RAPS sensitivity analyses did not suggest bias due to weak instruments. For the bidirectional analyses we did not observe significant associations between NHL and the risk of any of the autoimmune diseases (Supplementary Table 2).”  See Figure 2 |
|  | b) | Report results from other sensitivity analyses or additional analyses | 14-15 | “In the secondary analyses, we evaluated the autoimmune diseases that were at least nominally significantly associated with risk of NHL, i.e., T1D and sarcoidosis. In the analyses restricted to non-immune pathways, we observed ORs of 0.96 (95% CI: 0.87 to 1.07) for T1D and 0.96 (95% CI: 0.88 to 1.06) for sarcoidosis, respectively, supporting the main analyses.  Figure 3 shows the IVW analyses of genetically-predicted susceptibility to T1D and sarcoidosis, and the risk of NHL subtypes. For T1D, the association with composite NHL appeared to be driven by follicular lymphoma, with an OR of 0.91 (95% CI: 0.86 to 0.96, P = 1 ×10-3). Sarcoidosis was most strongly associated with other and unspecified types of NHL, with an OR of 0.86 (95% CI: 0.75 to 0.97, P = 1.8×10-2).” |
|  | c) | Report any assessment of direction of causal relationship (e.g., bidirectional MR) | 14 | “For the bidirectional analyses we did not observe significant associations between NHL and the risk of any of the autoimmune diseases (Supplementary Table 2).” |
|  | d) | When relevant, report and compare with estimates from non-MR analyses | 17 | “In particular, according to an umbrella review of meta-analyses, the summary relative risks for the associations between T1D and NHL and sarcoidosis and NHL were 1.55 (95% CI: 1.15 to 2.08) and 1.43 (95% CI: 1.03 to 1.99), respectively.” |
|  | e) | Consider additional plots to visualize results (e.g., leave-one-out analyses) | Not applicable |  |
|  | **DISCUSSION** |  |  |  |
| 14 | **Key results** | Summarize key results with reference to study objectives | 16 | “In this MR study of ten autoimmune diseases previously linked to an increased risk of NHL, we found that genetically predicted susceptibility to T1D, and to some extent, genetically predicted susceptibility to sarcoidosis, were associated with a reduced risk of NHL. While these findings were consistent across a wide range of sensitivity analyses, no clear associations were observed between the other eight autoimmune diseases and risk of NHL. Using an approach that attempts to address potential residual confounding and reverse causation, our findings contradict those reported in previous traditional observational studies. This highlights the need for future studies with different datasets, approaches, and populations to further examine the potential associations between these autoimmune diseases and the risk of NHL.” |
| 15 | **Limitations** | Discuss limitations of the study, taking into account the validity of the IV assumptions, other sources of potential bias, and imprecision. Discuss both direction and magnitude of any potential bias and any efforts to address them | 19 | The entire paragraph of “Our study had several limitations……” |
| 16 | **Interpretation** |  |  |  |
|  | a) | Meaning: Give a cautious overall interpretation of results in the context of their limitations and in comparison with other studies | 16-17 | The entire paragraphs of Principal findings and Context of primary findings |
|  | b) | Mechanism: Discuss underlying biological mechanisms that could drive a potential causal relationship between the investigated exposure and the outcome, and whether the gene-environment equivalence assumption is reasonable. Use causal language carefully, clarifying that IV estimates may provide causal effects only under certain assumptions | 18 | “Fourth, it is unclear whether the increased NHL risk is attributable to the autoimmune diseases or the immunosuppressants that are used as treatments.47,52 Given that studies have suggested that immunosuppressive patients (e.g., patients with HIV infection or an organ transplantation) have an increased risk of developing NHL,53 it is possible that autoimmune diseases may have a limited impact on the increased NHL risk. Fifth, it has been suggested that the pathways involved in autoimmune disease and cancer development may work in opposite directions,54 meaning the increased risk of autoimmune diseases can reduce the susceptibility to NHL.” |
|  | c) | Clinical relevance: Discuss whether the results have clinical or public policy relevance, and to what extent they inform effect sizes of possible interventions | 19-20 | “While our study addressed potential residual confounding that may have been present in previous observational studies, the MR design may be biased if the instrumental variable assumptions do not hold. Thus, the association between autoimmune diseases and risk of NHL needs further investigation with evidence triangulation using different datasets, populations, and approaches.” |
| 17 | **Generalizability** | Discuss the generalizability of the study results (a) to other populations, (b) across other exposure periods/timings, and (c) across other levels of exposure | 19 | “…this study is limited to populations of European ancestry, which have the highest incidence of NHL, and it is unclear whether the findings can be generalised beyond this population.” |
|  | **OTHER INFORMATION** |  |  |  |
| 18 | **Funding** | Describe sources of funding and the role of funders in the present study and, if applicable, sources of funding for the databases and original study or studies on which the present study is based | 21 | “Dr. Rogne received funding from CTSA Grant Number UL1 TR001863 from the National Center for Advancing Translational Science (NCATS), a component of the NIH. The contents of this manuscript are solely the responsibility of the authors and do not necessarily represent the official views of NIH. The NIH had no role in the design and conduct of the study; collection, management, analysis, and interpretation of the data; preparation, review, or approval of the manuscript; and decision to submit the manuscript for publication.” |
| 19 | **Data and data sharing** | Provide the data used to perform all analyses or report where and how the data can be accessed, and reference these sources in the article. Provide the statistical code needed to reproduce the results in the article, or report whether the code is publicly accessible and if so, where | 22 | “The data will be made available via a publicly accessible repository on publication. The code for analyses is included as supplementary information.” |
| 20 | **Conflicts of Interest** | All authors should declare all potential conflicts of interest | 21-22 | “All authors have completed the ICMJE uniform disclosure form at www.icmje.org/coi_disclosure.pdf and declare: In the past 36 months, Dr. Wallach reported receiving grant support from the FDA, Arnold Ventures, Johnson & Johnson through Yale University, and the National Institute on Alcohol Abuse and Alcoholism of the National Institutes of Health (NIH) under award 1K01AA028258; serving as a consultant for Hagens Berman Sobol Shapiro LLP and Dugan Law Firm APLC; and serving as a medRxiv affiliate. Dr. Ma received research funding from the NIH and the Frederick A. DeLuca Foundation and served as a consultant for Bristol Myers Squibb.” |

This checklist is copyrighted by the Equator Network under the Creative Commons Attribution 3.0 Unported (CC BY 3.0) license.

1.	Skrivankova VW, Richmond RC, Woolf BAR, Yarmolinsky J, Davies NM, Swanson SA, et al. Strengthening the Reporting of Observational Studies in Epidemiology using Mendelian Randomization (STROBE-MR) Statement. JAMA. 2021;under review.

2.	Skrivankova VW, Richmond RC, Woolf BAR, Davies NM, Swanson SA, VanderWeele TJ, et al. Strengthening the Reporting of Observational Studies in Epidemiology using Mendelian Randomisation (STROBE-MR): Explanation and Elaboration. BMJ. 2021;375:n2233.
